# Effects of Fingertip Vibrotactile Stimulation on Postural Control in Community-Dwelling Older Adults: A Comparison Across Age Groups

**DOI:** 10.64898/2026.08.17.26360566

**Authors:** Takato Nishida, Shin Murata, Ryosuke Yamamoto, Shun Sawai, Shoya Fujikawa, Yusuke Shizuka, Naoki Shimizu, Koji Shimatani, Keisuke Shima, Hideki Nakano

## Abstract

Age-related decline in postural control is an important factor that increases the fall risk of older adults. Fingertip vibrotactile stimulation has been developed to provide light touch-like somatosensory input. However, evidence regarding differences among older age groups is limited. This study examined the effects of fingertip vibrotactile stimulation on postural control in 348 community-dwelling older adults classified as young-old (age 65–74 years), old-old (age 75–84 years), and oldest-old (age 85 years or older). Participants stood with eyes closed and feet together under stimulation and no stimulation conditions. The center of pressure (COP) velocity and COP area were measured using a force plate. The natural log-transformed COP area was used for the analysis. Linear mixed models were used to examine the effects of age group, stimulation conditions, and measurement segments. The COP velocity under the stimulation condition was significantly lower than that under the no stimulation condition; however, the COP area did not change significantly. Significant main effects of age group were observed for both COP indices, but no interaction between age group and stimulation condition was observed. Fingertip vibrotactile stimulation may reduce the COP velocity across older age groups, thus reflecting the effects on postural adjustment frequency.

## Introduction

As the worldwide population ages, fall prevention and independent living support have become important public health and social issues. Older adults need to maintain and improve their physical function to continue living independently. Standing postural control is closely associated with the fall risk, and age-related decline in standing postural control is considered a major factor that contributes to falls [1]. Therefore, investigating strategies to maintain and improve postural control is important to extending the healthy life expectancy.

Light touch (LT) contact is a known method of stabilizing the standing posture. Merely touching a support surface lightly can reduce postural sway, even without mechanical support [2]. This effect is thought to result from the use of somatosensory information, such as tactile and proprioceptive information, for postural control [3,4]. Furthermore, electroencephalographic studies have shown that under LT conditions, activity changes occur in sensory integration-related regions, mainly in the parietal lobe, thereby improving the accuracy of body position estimation [5]. Additionally, increased somatosensory input may reorganize the activity of cortical networks involved in postural control, thereby improving postural stability even in the absence of mechanical support. These findings indicate that LT involves a mechanism that improves postural control through sensory integration processes in the central nervous system.

However, compared with younger adults, older adults exhibit greater contact force under LT conditions [6], suggesting limitations in the direct application of conventional LT methods among older adults. Although the sway-reducing effect of LT on older adults has been reported, it is unclear whether responses to LT differ among age groups within the older adult population because of age-related declines in sensory integration and physical function.

Recently, StA²BLE (UNTRACKED Inc., Yokohama, Japan), which is a device that provides LT-like somatosensory input using vibrotactile stimulation, has been developed to address this issue. StA²BLE (UNTRACKED Inc.) is a wearable device that estimates virtual contact force based on acceleration information from the fingertip and provides real-time vibrotactile feedback according to the magnitude of the estimated force, thereby delivering LT-like tactile somatosensory input even without actual contact with a support surface. Shima et al. reported that this method produces postural stabilization effects comparable to those of conventional LT [7], suggesting the possibility of providing stable sensory input even for older adults.

However, few studies have examined the effects of somatosensory input provided by vibrotactile stimulation on postural control in older adults [8]. Furthermore, because physical function varies substantially with advancing age, even within the older adult population, whether consistent effects can be obtained across age groups ranging from young-old to oldest-old adults is unclear.

Therefore, this study aimed to clarify the effects of somatosensory input provided by fingertip vibrotactile stimulation on postural control in community-dwelling older adults across different age groups.

## Results

### Participant characteristics

The age, sex, Mini-Mental State Examination (MMSE) score, knee extension strength, and toe grip strength of 348 participants included in the analysis are shown in Table 1. Between-group comparisons revealed significant differences in sex (P = 0.007), MMSE score (P = 0.004), knee extension strength (P < 0.001), and toe grip strength (P < 0.001). Post hoc comparisons showed that the number of male participants in the old-old group (age 75–84 years; P = 0.024) and oldest-old group (age 85 years or older; P = 0.012) was significantly higher than that in the young-old group (age 65–74 years). The MMSE scores in the oldest-old group were significantly lower than those in the young-old group (P = 0.003). Knee extension strength in the old-old group (P = 0.043) and oldest-old group (P = 0.001) was significantly lower than that in the young-old group. Similarly, toe grip strength in the old-old group (P = 0.005) and oldest-old group (P < 0.001) was significantly lower than that in the young-old group.

**Table 1.**
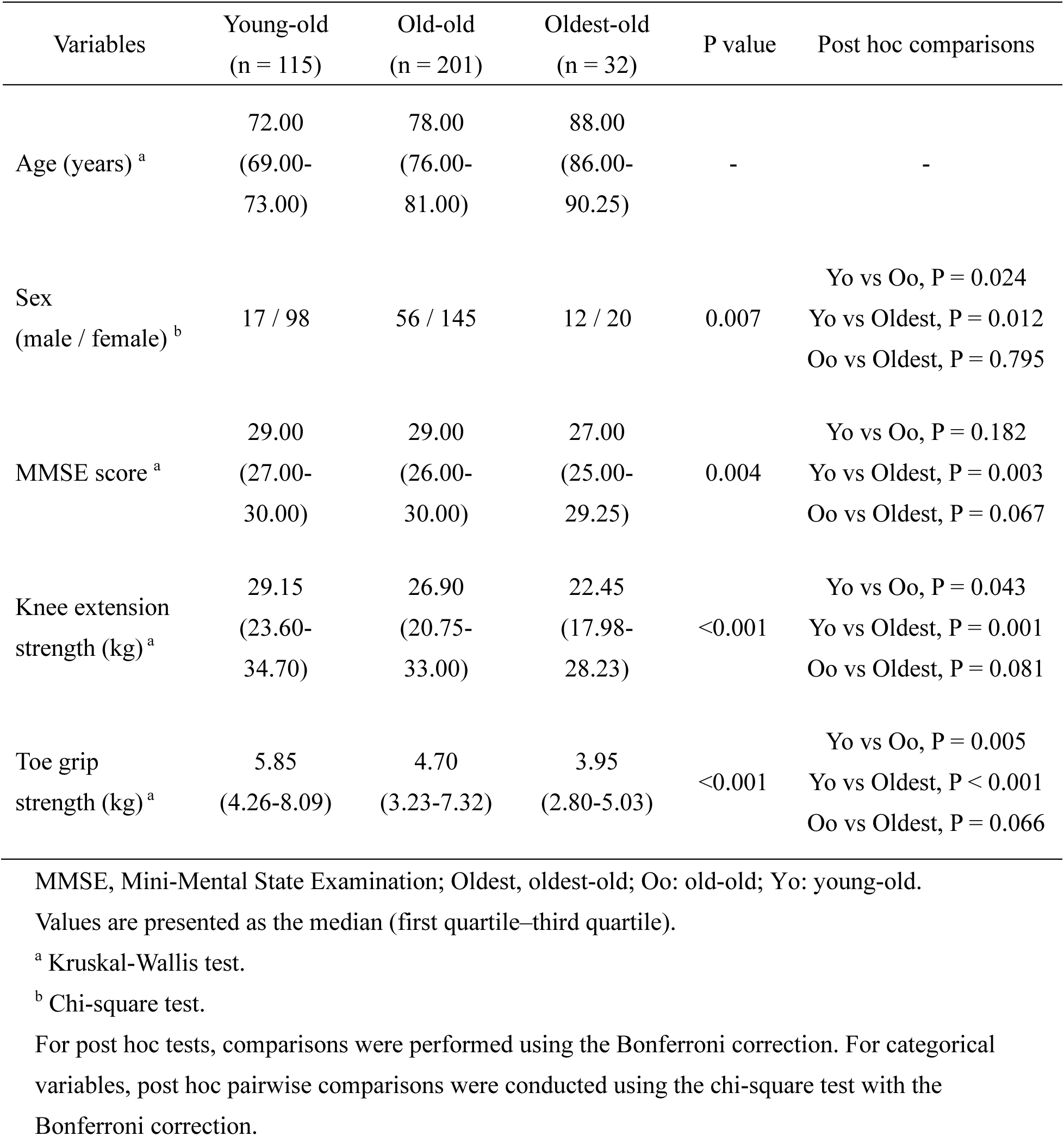
Participant characteristics.

### COP velocity

The means and standard deviations of the COP velocity in each group with each condition are shown in Table 2. The COP velocity increased with older age. The linear mixed model showed a significant main effect of group (F (2, 345) = 13.28, P < 0.001; partial η² = 0.07) (Table 3). Multiple comparisons with the Bonferroni correction showed that values in the young-old group were significantly lower than those in the old-old group (P < 0.001) and oldest-old group (P < 0.001). In contrast, no significant difference between the old-old and oldest-old groups was observed (P = 0.060) (Figure 1). The main effect of stimulation condition was also significant. The COP velocity under the stimulation condition was significantly lower than that under the no stimulation condition (F (1, 1035) = 8.34; P = 0.004; partial η² = 0.01) (Figure 1, Table 3). In contrast, no significant main effect of measurement segment was observed (F (1, 1035) = 0.69; P = 0.405; partial η² < 0.01). Furthermore, group–stimulation condition (F (2, 1035) = 0.58; P = 0.561; partial η² < 0.01), group–measurement segment (F (2, 1035) = 0.72; P = 0.486; partial η² < 0.01), stimulation condition–measurement segment (F (1, 1035) = 0.42; P = 0.516; partial η² < 0.01), and group–stimulation condition–measurement segment (F (2, 1035) = 0.02; P = 0.978; partial η² < 0.01) interactions were not significant (Table 3).

**Fig. 1.**
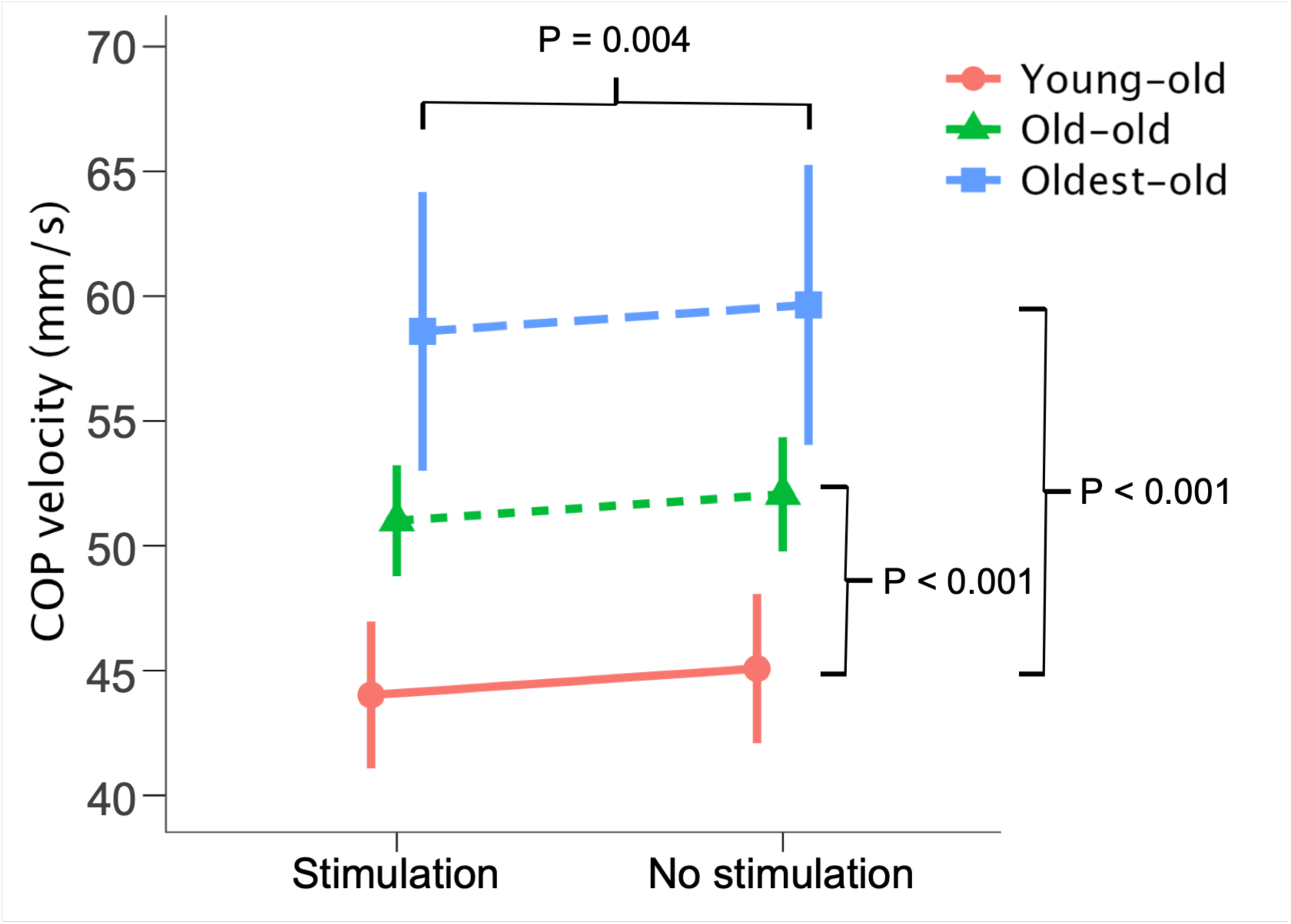
Comparison of the COP velocity under stimulation and no stimulation conditions across age groups. The COP velocity under the stimulation condition was significantly lower than that under the no stimulation condition. The COP velocity in the young-old group was significantly lower than that in the old-old and oldest-old groups. Data are presented as the estimated marginal means, and error bars indicate standard errors. The horizontal bracket indicates the main effect of stimulation condition, and the vertical brackets indicate significant post hoc comparisons with the Bonferroni correction among age groups. COP, center of pressure.

**Table 2.** Center of pressure velocity under each condition (mm/s)

| Group | Stimulation |  | No stimulation |  |
| --- | --- | --- | --- | --- |
|  | First segment | Second segment | First segment | Second segment |
| Young-old<br>(n = 115) | 43.53 ± 12.06 | 44.22 ± 13.24 | 44.39 ± 13.34 | 44.73 ± 13.49 |
| Old-old<br>(n = 201) | 51.02 ± 17.99 | 51.18 ± 17.97 | 52.59 ± 19.44 | 52.18 ± 18.31 |
| Oldest-old<br>(n = 32) | 58.19 ± 19.79 | 58.83 ± 15.63 | 59.33 ± 17.63 | 59.58 ± 18.20 |

Values are presented as the mean ± standard deviation.

**Table 3.** Results of the linear mixed model of the center of pressure velocity.

| Parameter | F (df 1, df 2) | P value | Partial $\eta^2$ |
| --- | --- | --- | --- |
| Group | 13.28 (2, 345) | <0.001 | 0.07 |
| Stimulation | 8.34 (1, 1035) | 0.004 | 0.01 |
| Segment | 0.69 (1, 1035) | 0.405 | <0.01 |
| Group–stimulation interaction | 0.58 (2, 1035) | 0.561 | <0.01 |
| Group–segment interaction | 0.72 (2, 1035) | 0.486 | <0.01 |
| Stimulation–segment interaction | 0.42 (1, 1035) | 0.516 | <0.01 |
| Group–stimulation–segment interaction | 0.02 (2, 1035) | 0.978 | <0.01 |

df 1, numerator degrees of freedom; df 2, denominator degrees of freedom; partial $\eta^2$ , partial eta squared.

### Natural log-transformed COP area

The means and standard deviations of the natural log-transformed COP area in each group and with each condition are shown in Table 4. Similar to the COP velocity, the COP area increased with older age. The linear mixed model showed a significant main effect of group (F (2, 345) = 10.50; P < 0.001; partial η² = 0.06) (Table 5). Multiple comparisons with the Bonferroni correction showed that values in the young-old group were significantly lower than those in the old-old group (P = 0.001) and oldest-old group (P < 0.001). No significant difference was observed between the old-old and oldest-old groups (P = 0.128) (Figure 2). In contrast, no significant main effect of stimulation condition was observed (F (1, 1035) = 0.01; P = 0.921; partial η² < 0.01). Additionally, the main effect of measurement segment was not significant (F (1, 1035) = 2.58; P = 0.108; partial η² < 0.01) (Table 5). Furthermore, group–stimulation condition (F (2, 1035) = 0.95; P = 0.385; partial η² < 0.01), group–measurement segment (F (2, 1035) = 0.44; P = 0.643; partial η² < 0.01), stimulation condition–measurement segment (F (1, 1035) = 3.18; P = 0.075; partial η² < 0.01), and group–stimulation condition– measurement segment (F (2, 1035) = 1.16; P = 0.314; partial η² < 0.01) interactions were not significant (Table 5).

**Fig. 2.**
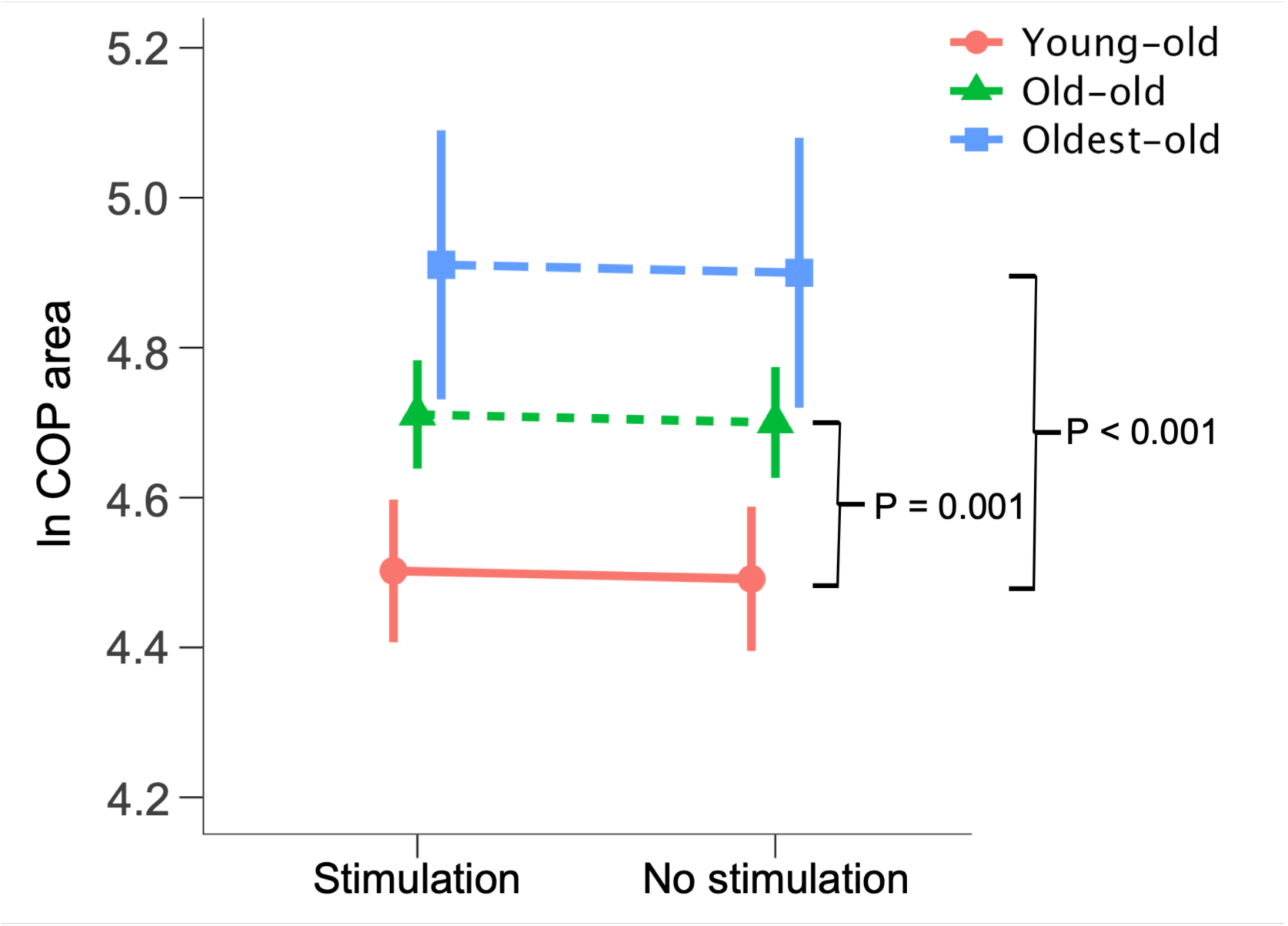
Comparison of the natural log-transformed COP area under stimulation and no stimulation conditions across age groups. The COP area was measured in mm² and natural log-transformed before the analysis. The natural log-transformed COP area in the young-old group was significantly lower than that in the old-old and oldest-old groups. No significant main effect of stimulation condition or group–stimulation condition interaction was observed. Data are presented as estimated marginal means, and error bars indicate standard errors. The vertical brackets indicate significant post hoc comparisons with the Bonferroni correction among age groups. COP, center of pressure.

**Table 4.** Natural log-transformed center of pressure area under each condition Group.

| Group | Stimulation |  | No stimulation |  |
| --- | --- | --- | --- | --- |
|  | First segment | Second segment | First segment | Second segment |
| Young-old<br>(n = 115) | 4.54 ± 0.56 | 4.49 ± 0.54 | 4.50 ± 0.60 | 4.45 ± 0.58 |
| Old-old<br>(n = 201) | 4.73 ± 0.59 | 4.69 ± 0.54 | 4.74 ± 0.63 | 4.67 ± 0.58 |
| Oldest-old<br>(n = 32) | 4.84 ± 0.52 | 4.94 ± 0.43 | 4.97 ± 0.42 | 4.88 ± 0.51 |

Values are presented as the mean ± standard deviation. The center of pressure area was measured in mm<sup>2</sup> and naturally log-transformed before the analysis.

**Table 5.** Results of the linear mixed model of the natural log-transformed center of pressure area.

| Parameter | F (df 1, df 2) | P value | Partial $\eta^2$ |
| --- | --- | --- | --- |
| Group | 10.50 (2, 345) | <0.001 | 0.06 |
| Stimulation | 0.01 (1, 1035) | 0.921 | <0.01 |
| Segment | 2.58 (1, 1035) | 0.108 | <0.01 |
| Group–stimulation interaction | 0.95 (2, 1035) | 0.385 | <0.01 |
| Group–segment interaction | 0.44 (2, 1035) | 0.643 | <0.01 |
| Stimulation–segment interaction | 3.18 (1, 1035) | 0.075 | <0.01 |
| Group–stimulation–segment interaction | 1.16 (2, 1035) | 0.314 | <0.01 |

df 1, numerator degrees of freedom; df 2, denominator degrees of freedom; partial $\eta^2$ , partial eta squared.

## Discussion

This study aimed to clarify how somatosensory input provided by fingertip vibrotactile stimulation affects postural control in older adults across different age groups. The results indicated that COP velocity significantly decreased with vibrotactile stimulation; however, no significant change in the COP area was observed. Additionally, the main effects of group were observed for both indices, but a group–stimulation condition interaction was not observed. These findings suggest that somatosensory input provided by vibrotactile stimulation has a certain effect on postural control in older adults and a limited effect on reducing the magnitude of postural sway, and that this effect may be common across age groups.

The decrease in the COP velocity indicated that vibrotactile stimulation affected the frequency of postural adjustments. The COP velocity reflects the amount of COP displacement per unit time, and an increase in COP velocity is considered to indicate more frequent postural adjustments [9,10]. In the present study, a main effect of stimulation condition was observed. The COP velocity under the stimulation condition was significantly lower than that under the no stimulation condition. Previous studies of LT have reported that weak somatosensory input from fingertip contact affects postural control. Changes in fingertip contact force can occur before lower limb muscle activity and can be reflected as changes in the COP approximately 300 ms later [11]. This temporal relationship suggests that somatosensory information is integrated as a long-loop reflex mediated by the cerebral cortex and predictively regulates postural muscle activity. Therefore, LT is considered to promote a predictive control mechanism, such as feedforward control, in which postural fluctuations are predicted based on externally obtained somatosensory information and postural control is performed in advance. Vibrotactile stimulation used in this study may have functioned as sensory input that substituted for or enhanced the somatosensory input provided by LT. Specifically, somatosensory information added by fingertip vibrotactile stimulation may have been integrated in the central nervous system and promoted predictive control of postural fluctuations, thereby suppressing unnecessary postural corrections and reducing fine reciprocal movements of the COP. Therefore, the decrease in the COP velocity induced by vibrotactile input may indicate a reduction in excessive postural corrections and a shift toward a more efficient and predictive postural control strategy.

In relation to the main research question of this study, the absence of a group–stimulation condition interaction was an important observation. Somatosensory information plays a central role in postural control; with aging, reliance on proprioception relatively increases as visual and vestibular functions decline [12]. Furthermore, the accuracy of sensory input integration declines with aging [13]. In contrast, the ability to reweight sensory information according to the reliability of each sensory input is preserved to some extent even in older adults [14]. These results also suggest that although older adults may rely on somatosensory information, the sensory reweighting ability may be preserved. Therefore, the somatosensory input added by vibrotactile stimulation may have been effectively used in all age groups and contributed to postural control, resulting in reduced COP velocity. Consequently, the effects of vibrotactile stimulation may not be limited to a specific age group and may occur commonly across older adults in different age groups. These results suggest that the manner in which somatosensory input provided by vibrotactile stimulation is used may not differ substantially among age groups; furthermore, at least within the population examined in this study, the responses to additional sensory input may be relatively uniform regardless of the age group.

In contrast, no main effect of vibrotactile stimulation on the COP area was observed. The COP area is widely used as a quantitative index that reflects the stability of static posture [1]. Previous LT studies that included younger and older adults have shown greater reductions in path length among older adults, whereas findings of the COP area have not been consistent; therefore, its effects are unclear [15]. Similarly, consistent with previous findings, no changes in the COP area were observed with vibrotactile stimulation in this study. Furthermore, in this study, lower limb muscle strength in the old-old and oldest-old groups was significantly lower than that in the young-old group. Lower limb muscle weakness is associated with an increased COP area [16], suggesting that physical function, particularly motor output capacity, is involved in the magnitude of postural sway. Thus, age-related declines in lower limb muscle strength may have limited the reduction in the COP area.

Collectively, these findings suggest that the provision of sensory input by vibrotactile stimulation affects qualitative aspects of postural control, such as the frequency of postural adjustments, whereas quantitative aspects, such as the magnitude of postural sway, may primarily depend on motor output capacity. Therefore, changes in sensory input alone may not have been sufficient to improve the COP area.

The finding that both the COP velocity and COP area in the old-old and oldest-old groups were higher than those in the young-old group as main effects of group was considered to reflect age-related decline in the postural control ability. With aging, the postural control system becomes vulnerable because of declining motor output, such as muscle weakness, declining integration of visual and somatosensory information, and delayed postural responses [13]. In the present study, both the COP velocity and COP area increased with age, suggesting that age-related effects were reflected in the COP indices.

Additionally, no main effect of measurement segment was observed for the COP velocity or COP area, indicating that no consistent changes between measurement segments were identified, at least under the measurement conditions used in this study.

This study had several limitations. First, the number of participants in the oldest-old group was smaller than that in the young-old and old-old groups, which limited the statistical power to detect differences in stimulation effects among age groups. Second, because electromyography, brain activity measurements, and sensory function tests were not performed in this study, we could not directly determine whether the decrease in the COP velocity was attributable to sensory integration, predictive control, or motor output. Third, the participants in this study were community-dwelling older adults with relatively high levels of independence; therefore, caution is needed when generalizing the findings to older adults who require long-term care or populations at high risk for falls.

This study showed that somatosensory input provided by fingertip vibrotactile stimulation may reduce COP velocity and improve aspects related to the frequency of postural adjustments in community-dwelling older adults. In contrast, no significant change was observed in the COP area, indicating that the effect on the magnitude of postural sway was limited. Additionally, because no age group–vibrotactile stimulation interaction was observed, the effects of fingertip vibrotactile stimulation may be generally consistent across age groups of younger and older adults.

## Methods

### Study design

This cross-sectional study of community-dwelling older adults included a repeated-measures design to compare changes in postural control indices with and without fingertip vibrotactile stimulation within the same participants.

### Participants

The participants were 382 community-dwelling older adults who participated in physical fitness measurement events held in Imari City, Saga Prefecture, between August 18 and 21, 2025, and Yasu City, Shiga Prefecture, between September 1 and 5, 2025. The exclusion criteria were as follows: age younger than 65 years; MMSE score <24; difficulty performing the measurement task; and did not provide consent to participate in this study. A total of 348 participants who did not meet any of these exclusion criteria were included in the analysis and classified into the young-old (age 65–74 years), old-old (age 75–84 years), and oldest-old (age 85 years and older) groups (Figure 3).

**Fig. 3.**
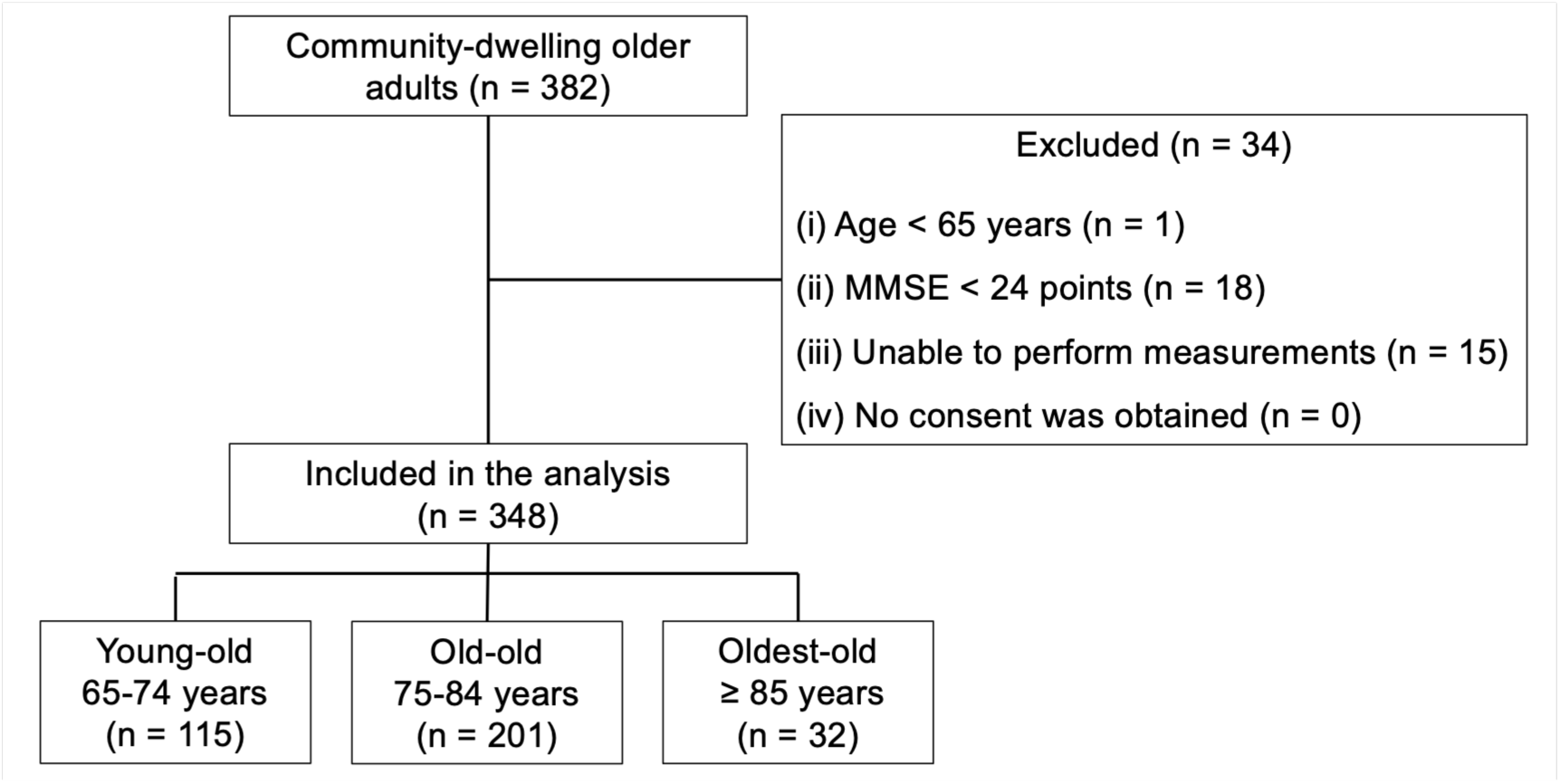
Participant selection flowchart. A total of 382 community-dwelling older adults were screened. After applying the exclusion criteria, 348 participants were included in the analysis and classified into the young-old, old-old, and oldest-old groups. MMSE, Mini-Mental State Examination.

### Ethics

This study was conducted in accordance with the Declaration of Helsinki and approved by the Research Ethics Committee of Kyoto Tachibana University (approval number: 25-36). All procedures were performed in accordance with relevant guidelines and regulations. All participants received written and oral explanations of the purpose and contents of the study in advance, and written informed consent was obtained from each participant.

### Measures

Age and sex were collected as participant characteristics. Knee extension strength and toe grip strength were measured to assess lower limb muscle strength. Knee extension strength was measured using a single-leg muscle strength measurement platform (TKK 5715; Takei Scientific Instruments Co., Ltd., Niigata, Japan) [17]. Participants were seated with the knee joint flexed at 90°, and a belt connected to a tension meter (TKK 5710; Takei Scientific Instruments Co., Ltd., Niigata, Japan) was attached to the ankle. Participants were instructed to extend the knee joint with maximal effort. Measurements were performed twice on each side, and the mean of the maximum values obtained from the right and left sides was used as the representative value.

Toe grip strength was measured using a toe grip dynamometer (TKK 3362; Takei Scientific Instruments Co., Ltd.) [18]. Participants were seated at the edge of a chair with the upper limbs crossed in front of the chest, hip and knee joints flexed at 90°, and ankle joint in a neutral position between plantarflexion and dorsiflexion. The handle of the dynamometer was positioned at the first metatarsophalangeal joint of the dominant leg. After sufficient practice, measurements were performed twice with rest intervals, and the maximum value was used as the representative value. In this study, the dominant leg was defined as the leg used to kick a ball.

#### Postural control ability

To examine changes in postural control induced by somatosensory input, vibrotactile stimulation was applied to the pulp of the index finger of the dominant hand using StA²BLE (UNTRACKED Inc.), and changes in the COP with and without stimulation were measured. Hand dominance was determined by self-report. The COP was measured using a force plate (Wii Balance Board; Nintendo Co., Ltd., Kyoto, Japan) [19]. Participants maintained a standing position for 60 seconds with their eyes closed and feet together. During the measurement, participants were instructed to repeatedly perform flexion and extension movements of the elbow joint on the side of the dominant hand at a constant rhythm to generate vibrotactile feedback from StA²BLE (UNTRACKED Inc.). The movement rate was set at 120 bpm.

During the measurement, vibrotactile stimulation was applied to the index finger in two segments for each condition as follows: stimulation, no stimulation, stimulation, and no stimulation. The duration of each segment was randomized for each participant and was within the range of 10 to 20 seconds. The total duration of each condition was set to 30 seconds. Postural sway data were sampled at 100 Hz, and the COP velocity (mm/s) and standard deviation area (COP area; mm²) were calculated as COP indices. Because the COP area showed a right-skewed distribution and heterogeneity of variance, the natural log-transformed value was used in the analysis to satisfy the assumption of normality. The COP velocity was calculated by dividing the COP path length in each segment by the measurement time of that segment.

#### Cognitive function

Cognitive function was assessed using the MMSE. The MMSE is widely used as an index of global cognitive function and consists of 11 items [20]. In this study, the MMSE was used as a descriptive index, and a score <24 was used as an exclusion criterion.

### Statistical analysis

The normality of each measurement variable was examined using the Shapiro–Wilk test. All analyses included 348 participants: 115 young-old adults, 201 old-old adults, and 32 oldest-old adults. Between-group comparisons of the MMSE score, knee extension strength, and toe grip strength were performed using the Kruskal–Wallis test because these variables did not meet the assumption of normality. Sex was compared among age groups using the chi-square test. For variables with significant between-group differences, post hoc pairwise comparisons were performed using the Bonferroni correction, and Bonferroni-adjusted P values are reported.

The COP velocity and natural log-transformed COP area were analyzed as dependent variables using linear mixed models. Linear mixed models were used because COP indices were measured repeatedly within the same participants under stimulation and no stimulation conditions. Group (young-old, old-old, and oldest-old), stimulation condition (stimulation and no stimulation), and measurement segment (first segment and second segment) were entered as fixed effects. The group–stimulation condition, group–measurement segment, stimulation condition–measurement segment, and group–stimulation condition–measurement segment interactions were also examined. The participant identification number was entered as a random effect to account for repeated measurements within participants. There were no missing data for the COP variables. Each participant contributed four segment-level observations, consisting of two stimulation segments and two no stimulation segments; therefore, each linear mixed model included 1,392 observations.

The assumptions of the linear mixed models were assessed by inspecting residual Q–Q plots and residual-versus-fitted plots. Because the raw COP area showed a right-skewed distribution and heterogeneity of variance, the natural log-transformed COP area was used in the analysis. When significant main effects or interactions were observed, tests of simple main effects and multiple comparisons using the Bonferroni method were performed. Partial η² was calculated as the effect size.

All statistical analyses were performed using SPSS Statistics version 30 (IBM Corp., Armonk, NY, USA). All statistical tests were two-sided when applicable. The significance level was set at α = 0.05. Exact P values were reported whenever possible, and P < 0.001 was reported as P < 0.001.

## Data Availability

The datasets generated and/or analyzed during the current study are not publicly available because of privacy and ethical restrictions but are available from the corresponding author on reasonable request and with permission from the relevant ethics committee, where applicable.

## Acknowledgments

The authors express their sincere gratitude to all participants in the physical fitness testing event and the staff for their cooperation.

## Author contributions

Conceptualization: TN and HN; Data curation: TN and HN; Formal analysis: TN and HN; Funding acquisition: TN, SM, and HN; Investigation: TN, SM, SS, SF, YS, NS, and HN; Methodology: TN and HN; Project administration: SM and HN; Resources: SM and HN; Software: K Shimatani and K Shima; Supervision: HN; Validation: TN, RY, SS, SF, YS, NS, and HN; Visualization: TN and HN; Writing – original draft: TN and HN; Writing – review & editing: TN, SM, RY, SS, SF, YS, NS, K Shimatani, K Shima, and HN.

